# Successful suppression of a field population of *Ae. aegypti* mosquitoes using a novel biological vector control strategy is associated with significantly lower incidence of dengue

**DOI:** 10.1101/19010678

**Authors:** Lisiane C Poncio, Filipe A dos Anjos, Deborah A de Oliveira, Débora Rebechi, Rodrigo N de Oliveira, Rodrigo F Chitolina, Marise L Fermino, Luciano G Bernardes, Danton Guimarães, Pedro A Lemos, Marcelo N E Silva, Rodrigo G M Silvestre, Emerson S Bernardes, Nitzan Paldi

## Abstract

**Background:** Despite extensive efforts to prevent recurrent *Aedes*-borne arbovirus epidemics, there is a steady rise in their global incidence. Vaccines/treatments show very limited efficacy and together with the emergence of mosquito resistance to insecticides, it has become urgent to develop alternative solutions for efficient, sustainable and environmentally benign mosquito vector control. Here we present a new Sterile Insect Technology (SIT)-based program that uses large-scale releases of sterile male mosquitoes produced by a highly effective, safe and environmentally benign method.

**Methods and findings:** To test the efficacy of this approach, a field trial was conducted in a Brazilian city (Jacarezinho), which presented a history of 3 epidemics of dengue in the past decade. Sterile male mosquitoes were produced from a locally acquired *Aedes aegypti* colony, and releases were carried out on a weekly basis for seven months in a predefined area. This treated area was matched to a control area, in terms of size, layout, historic mosquito infestation index, socioeconomic patterns and comparable prevalence of dengue cases in past outbreaks. Releases of sterile male mosquitoes resulted in up to 91.4% reduction of live progeny of field *Ae. aegypti* mosquitoes recorded over time. The reduction in the mosquito population was corroborated by the standard monitoring system (LIRAa index) as determined by the local municipality, which found that our treated neighborhoods were almost free of *Ae. aegypti* mosquitoes after 5 months of release, whereas neighborhoods adjacent to the treated area and the control neighborhoods were highly infested. Importantly, when a dengue outbreak started in Jacarezinho in March 2019, the effective mosquito population suppression was shown to be associated with a far lower incidence of dengue in the treated area (16 cases corresponding to 264 cases per 100,000 inhabitants) almost 16 times lower than the dengue incidence in the control area (198 cases corresponding to 4,360 dengue cases per 100,000 inhabitants).

**Conclusions:** Our data present the first demonstration that a SIT-based intervention has the potential to prevent the spread of dengue, opening exciting new opportunities for preventing mosquito-borne disease.

## Introduction

Mosquito-transmitted arboviruses are the cause of substantial human mortality and morbidity. Dengue is endemic in more than 100 countries in Africa, the Americas, the Eastern Mediterranean, Southeast Asia and the Western Pacific [1]. An estimated 500,000 people with severe dengue require hospitalization each year, a large proportion of whom are children [2]. The symptoms of dengue include high fever, severe headaches, muscle and joint pain, nausea, vomiting, swollen glands or rash [3]. Dengue itself is rarely fatal, but severe dengue is a potentially fatal complication, with symptoms including low temperature, severe abdominal pains, rapid breathing, bleeding gums and blood in vomit [2].

Although the World Health Organization (WHO) has announced its intention to target reduction of global incidence of dengue by 75% in the next decade [2], the apparent reality seems to be heading in the opposite direction [4]. Indeed, hyper-urbanization and climate change are driving the expanded range of the primary mosquito vector of dengue and other arboviruses [5–7]. Consequently, the high morbidity and consequent economic and resource burden on health services in endemic settings is substantial and increasing [1].

Since specific vaccines to arboviruses have been presenting very limited efficacy and no treatments are available other than management [8], the main strategy to prevent the outbreak of epidemics still remains the control of the mosquito vectors, with the *Ae. aegypti* mosquito being by far the most common and efficient vector of dengue [9]. However, the traditional methods of vector control, such as the mechanical removal of potential breeding sites for mosquitoes and the use of insecticides [10], have been shown to be insufficient to prevent disease outbreaks, as has been evidenced by the continued rise of dengue incidence [4]. This has created an urgent need for alternative solutions to vector control.

Several biological paradigms for efficient *Ae. aegypti* control have created an interest both in the scientific and public-health communities over the past few years. One of these is the Sterile Insect Technique (SIT), which is based on the massive and continuous release of sterile male mosquitoes that mate with the wild females. Subsequently, these females do not generate viable offspring, which results in the gradual reduction of the local mosquito population [11]. As a method of insect control, SIT has several fundamental advantages, the most important of which is that, by definition, it provides species-level specificity, with no off-target effects [12]. In addition, another advantage of SIT programs is that there is virtually no risk of selecting resistant mosquito populations, which is one of the main criticisms related to chemical control (insecticides) [10].

Several SIT-based vector programs have already been shown to suppress mosquito populations in field studies [13–15]. Although SIT programs for vector control are well-known in the scientific community for many years [11], there are several fundamental limitations that preclude their wide implementation. The sterilization of mosquitoes by irradiation, for example, decreases competitiveness capacity of male mosquitoes [11]. The use of genetically modified (GM) mosquitoes is often shunned by the population and governments, due in part to fear of gene flow from released GM mosquitoes into the local gene pool [16]. Also, the technique based on cytoplasmic incompatibility (Incompatible Insect Technology – IIT), which utilizes mosquitoes infected with *Wolbachia* bacteria, seems to present several vulnerabilities, including the reduced competitiveness induced by some *Wolbachia* strains, risk of population replacement, selection of virus resistant to *Wolbachia*, and also limitations related to scaling up [13,17,18]. Above all, no such program has ever been able to directly demonstrate prevention of the spread of dengue or other arboviruses [13–15].

Herein we present a new and environmentally benign vector control intervention. This intervention comprised a series of actions implemented over one year in a Brazilian town (Jacarezinho) and included releases of sterile male mosquitoes (Natural Vector Control Mosquitoes-NVC) produced from the locally acquired *Ae. aegypti* mosquito population. The results of this intervention offered the first evidence that a SIT vector-control program can dramatically reduce the spread of dengue.

## Methods

### Establishment of local *Ae. aegypti* mosquito colony

*Ae. aegypti* mosquitoes from Jacarezinho were obtained via a field collection of eggs from the *Aedes* genus, using ovitraps distributed in several locations in the city of Jacarezinho in 2017. *Ae. aegypti* males and females from this F0 generation were mated and the females could individually lay their eggs. Then, all females that laid eggs from this F0 generation were collected and analyzed for the presence of dengue, Zika and chikungunya viruses, using a Real-time PCR method (Multiplex Dengue, Chikungunya, Zika virus, Genesig, USA). No infected females were detected and the eggs from these confirmed pathogen-free females were used to establish the mosquito colony of the Jacarezinho strain.

All the mosquitoes used in the study were reared in the Insectary of Forrest Brasil Tecnologia Ltda. located in Araucária city, in the state of Paraná, Brazil, and following all the biosafety parameters required for the process as defined by Environmental Institute of Paraná (IAP). The basic mosquito growth protocol and massive production of eggs was based on Rutledge et al. 1964 [19], with modifications. Briefly, freshly hatched larvae were placed in rearing trays and fed with commercially available alevin fish food (Supra Alevino), according to a predetermined regime to enable similar development for all production batches. Adult mosquitoes were fed on 10% sugar solution and kept at 26-28°C, 70-80% relative humidity and a 12:12 photoperiod. For massive production of eggs, adult females were mated with male mosquitoes (3 females: 1 male ratio) for three days and then fed with defibrinated sheep blood (Laborclin) using artificial feeders.

### Production of NVC sterile male mosquitoes

NVC mosquitoes were produced using a method that incorporates the use at the larval stage of specific dsRNAs, which targets the gene AAEL013723-PA, encoding a polypyrimidine tract binding protein (PTB) [20] and treatment with thiotepa at the pupal stage.

NVC sterile male mosquitoes used in all the experiments that were conducted under laboratory and semi-field conditions were produced as follows; the production process started by hatching *Ae. aegypti* eggs in 3 L dechlorinated water containing 150 mL of aged water and 0.15 mg fish food /ml at 26° - 26,5°C, for an overnight period. Subsequently, groups of one hundred first instar larvae were treated with a food formulation containing 1 mg of PTB-1 dsRNA (Table 1), 120 mg of yeast and 60 mg of fish food encapsulated in 1.2% (W/V) of sodium alginate particles. Larvae were fed with this formulation until they reached the third instar development phase. Then, third instar larvae were fed until the pupal stage with a second formulation, composed of 10 µg/mL of PTB-2 dsRNA (Table 1), 450 µL of PEG 20% (Polyethylene glycol 4000, Sigma), 200 µL of 0.01 g/mL Chitosan and 180 mg of Bovine Liver Powder encapsulated in 1.2 % (W/V) sodium alginate particles. To complete the sterilization process, a final step was performed after mechanical sorting of male and female pupae (Larval-Pupal Separator, Model 5412, John W. Hock Company, Florida, USA). Females were discarded, and males were kept in a 0.1% thiotepa solution, overnight, then extensively washed in acidified water (pH 3). Male pupae were finally placed in cages to emerge.

**Table 1.**
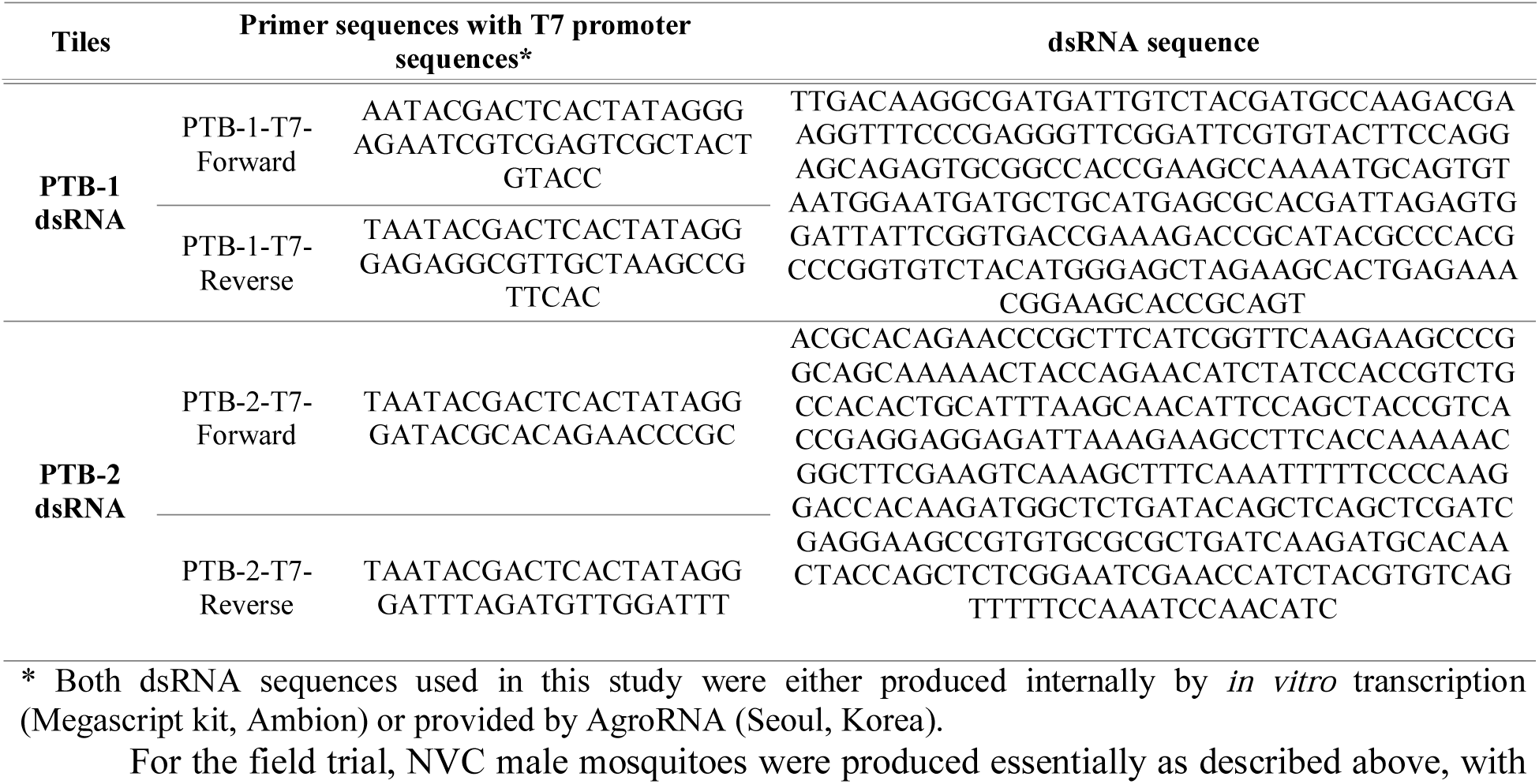
Primer sequences used to synthetize dsRNA sequences used to silence the PTB gene (AAEL013723-PA)

For the field trial, NVC male mosquitoes were produced essentially as described above, with several modifications necessary to adapt the protocol for large-scale production. These modifications included the higher number of larvae treated per batch and the reduction of dsRNA concentration during the first phase of NVC male mosquito production. In the pupae phase, males and females were mechanically sorted as described above, and all the batches of NVC production underwent a quality control to detect the potential presence of contaminating females. For this, a sample of at least 1,000 individuals was collected and analyzed under 10x magnification. A minimum threshold for pupal sorting accuracy of 99.8% males per group was imposed. All the batches of NVC pupae below this value was re-sorted until it was above 99.8%. Finally, approved batches of male pupae were subjected to thiotepa treatment. For large-scale production, the time of exposure of male pupae to thiotepa solution (at 0.6% W/V) was reduced to 2.5 hours, to avoid adult emergence during this step of treatment. Subsequently, thiotepa-treated NVC mosquitoes underwent three steps of washes to remove and inactivate any remnants of thiotepa solution that might stick to the outer surface of the pupae [21]: the first with tap water (for 10 min); the second with 0.0025 N H_2_SO_4_ solution at pH 2-3 (for 10 minutes) and the third and last one with 1 mm NaOH solution at pH 9 (for 20 minutes). This final step ensures complete degradation of thiotepa and derivatives into inert and non-toxic compounds [21]. Finally, pupae were rinsed in water to remove traces of the alkaline solution and then transferred to a container within the adult cage to emerge. After emerging as adults, NVC sterile male mosquitoes were fed 10% sugar solution.

### Fertility bioassay

To demonstrate the fertility status and competitiveness of NVC mosquitoes under laboratory conditions, NVC male mosquitoes (treated group) and untreated male mosquitoes (control group) were mated with virgin (fertile) females in the ratio of 1:3 (one male to three females). To test the competitiveness capacity of NVC mosquitoes, NVC males were allowed to mate with virgin females in the presence or absence of different proportions regular non-NVC males, according to the following experimental groups: fertile control (ten untreated virgin females and ten untreated males); sterile control (ten untreated virgin females and ten NVC treated mosquitoes); ratio 1:1 (ten untreated virgin females with five NVC and five untreated males) and ratio 10:1 (ten untreated virgin females with nine NVC and one untreated males). Three replicates from each group were prepared. Males and females were kept in the cage for a period of three to five days to allow mating.

Following the period for mating, females (from both fertility and competitiveness assays) were submitted to the steps of blood feeding, oviposition, embryonic development, hatching and counting. Details of the protocol can be found in the supplementary information (appendix S1).

### Semi-field trial

A semi-field experiment was carried out to test NVC male mosquitoes’ ability to compete with the wild male mosquitoes in ambient settings. The experiment was performed using a cage system that allowed mosquitoes to be exposed to local environmental conditions (temperature and humidity) but, at the same time, kept them in a biosafety environment so they would not be released into the wild. This experiment was performed in the city of Jacarezinho. Details of cage system can be found in appendix S2.

For the competitiveness assays, NVC mosquitoes were allowed to mate with virgin females inside the semi-field cages in the presence or absence of different proportions of normal males, according to the following experimental groups: fertile control (fertile males only); sterile control (sterile males only); ratio 1:1 (equal proportions of sterile and fertile males); ratio 10:1 (10 sterile males per 1 fertile male). The distribution of each of the groups of the experiment was performed in a random manner, using an Excel randomization worksheet. At the time of adult release, male mosquitoes were released before females. Only after all males were released, the females were released into the cages. Males and females were allowed to mate for a period of three days.

After the three-day mating period, a blood meal was provided for each semi-field cage. For this, three anesthetized mice were kept in each semi-field cage for a one-hour period, then all mice used were euthanized by anesthetic over-dose. All mice used in the experiments were provided by the Paraná Institute of Technology (TECPAR), after approval by the Local Ethics Committee (appendix S3). Three days after blood feeding, five ovitraps were placed inside each cage [22,23]. Females were allowed to oviposite for a period of five days, then, ovitraps were collected and transferred to the laboratory. Eggs were dried for five days to allow embryonic development and counted. Finally, eggs were hatched to verify the percentage of viable larvae. For this purpose, each paddle from each semi-field cage was kept in a tray with 2 L of water for 48 hours, then, hatched larvae were counted. The percentage of hatching was calculated based in the total number of eggs and viable larvae.

### Study location of field trial

Jacarezinho is a city in the northern region of Paraná state, located in southern Brazil (Figure 1A). Jacarezinho has a temperate climate, with well distributed rainfall throughout the year, and a mean annual precipitation of 1376 mm. The warmest and wettest month is January (average temperature of 25°C and 187 mm of precipitation), the coldest and driest month is June (average temperature of 17°C and 50 mm of precipitation) and the average temperature is 21°C. The study areas in Jacarezinho were chosen based on *Ae. aegypti* infestation rates and historical dengue epidemics, provided on a regular basis by the State Health Department of Paraná. Control and treated areas displayed similar number of inhabitants and socioeconomic characteristics, based on demographic data provided by the local municipality. Priority was given to neighborhoods with the highest historical rate of mosquito infestation and occurrence of dengue in past outbreaks. The control area was comprised of three neighborhoods which, together, correspond to an area of 81 hectares and have approximately 4,500 inhabitants. The treated area was also comprised of three neighborhoods with 77 hectares in total and approximately 6,000 inhabitants. As shown in Figure 1A, the areas chosen are separated by almost 4 km. To facilitate the releases of NVC mosquitoes and monitoring, the treated area was subdivided into three sub-regions (A, B and C) and microregions (Figure 1B).

**Figure 1.**
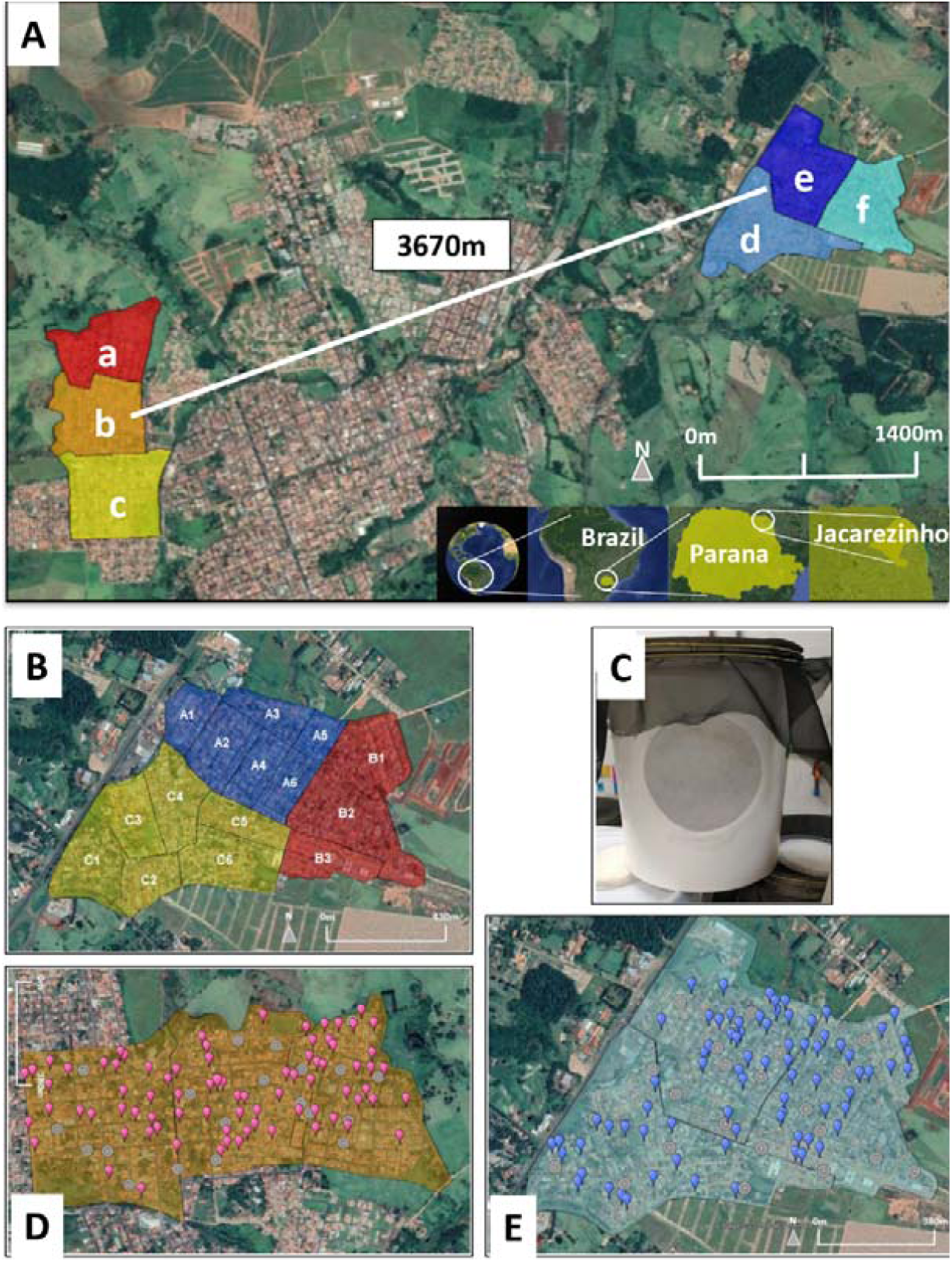
Satellite image (map data April 2019: Google, USA) showing the location of the study areas in Jacarezinho, South of Brazil. **A**. Map of Jacarezinho urban area and surroundings, showing the control and treated areas. The three neighborhoods chosen for the control Area (a-Dom Pedro Filipack, b-Vila Maria and c-Vila São Pedro) is shown on the left and neighborhoods chosen as the treated area (d-Vila Leão, e-Aeroporto and f-Novo Aeroporto) are shown on the right. Municipality of Jacarezinho, Paraná state, Brazil. **B**. The treated area was divided in microregions a, b and c. **C**. The cage where NVC mosquitoes were packed before releases. **D**. Map of control area showing the points where ovitraps (pink marks) were installed. **E**. Map of the treated area showing the location of the ovitraps (blue marks).

### NVC mosquito releases, *Ae. aegypti* surveillance and viable progeny

Seven-day-old NVC male mosquitoes were packed in cylindric plastic containers (4.2L of capacity, 210 mm height, 184 mm diameter, Figure 1C), at a maximum of 4,000 per container, and were fed with sugar solution until release. Mosquito releases were performed by manually opening the containers as a car goes through the streets of the treated area, according to the release schedule of the week. The number of NVC male mosquitoes released in each microregion were defined based on the monitoring of eggs collected in these areas in the previous week.

The monitoring of *Ae. aegypti* abundance was performed through the weekly installation of 100 ovitraps [22,23] in the houses or in the peridomiciliar area of the residences of both treated and control area. The selection of houses for the installation of ovitraps was performed in a random way. For this, all the houses from control and treated areas received numbering and a draw was performed, following the LIRAa methodology [24]. At least one resident of each house used for the traps signed a consent form that allowed ovitraps to be installed in their home (appendix S4). Ovitraps were installed and kept in the field for a period of 7 days, then removed to the laboratory and replaced with new ones. Once at the laboratory, eggs collected in each ovitrap were air dried for five days to complete the embryological development. Eggs were counted, and the hatching was performed by individually immersing each wooden paddle in a 0.0175% (W/V) solution of fish meal diluted in filtered water. The eggs were kept in this solution for 48 hours and then the larvae that hatched in this period were counted and considered viable progeny.

To monitor the abundance of *Aedes* genus population in Jacarezinho, prior to the period of NVC releases, **o**vitraps were installed in 52 points distributed throughout the town and replaced weekly. The eggs from each ovitrap were hatched together and larvae were reared to adulthood. Adult mosquitoes were then identified by species.

### Entomological and epidemiological data

Where indicated, official *Ae. aegypti* infestation data was provided by the Sanitary Surveillance of the Health Department of Jacarezinho according to *Ae. aegypti* Infestation Index Rapid Survey (LIRAa). This method was developed by the Brazilian Ministry of Health and consists of bimonthly larval sampling of *Ae. aegypti* in predetermined survey points, which are a function of the human population density and the number of existing buildings in the city [24]. LIRAa index is expressed as percentage of analyzed buildings where *Ae. aegypti* larvae are found. LIRAa index below 1% classify the area surveyed as satisfactory; LIRAa between 1% and 3.9%, the situation is defined as “alert” and LIRAa index above 4% indicates a risk of a dengue outbreak.

Epidemiological data regarding dengue cases in Jacarezinho was provided by the Epidemiological Surveillance of the Health Department of Jacarezinho. All the patients presenting dengue symptoms were reported to the local authority responsible. Subsequently, dengue rapid test (ELISA method, detection of NS1 using both IgM and IgG) was applied to each and every of these patients, to confirm (or not) the dengue diagnosis. In parallel, a blood sample from each patient was collected and analyzed by the Central Laboratory of Paraná State (LACEN), the regional authority, for further analysis using RT - PCR MULTIPLEX and dengue serological identification.

### Analysis of field population suppression and statistics

Estimation of field population suppression was performed as previously described in Gorman et al., 2015 [25], with modifications. Weekly moving averages relative to the same period at each control area were calculated according to the equation *M = (T*_*a*_*/C*_*a*_*)/(T*_*b*_*/C*_*b*_*) – 1*, where M is the population change, *T*_*a*_ is mean larvae per point in the treated area after release, *C*_*a*_ is mean larvae per point in control area after release; *T*_*b*_ is mean larvae per point in treated area before release and *C*_*b*_ is mean larvae per point in control area before release. This was done by comparing data obtained weekly against baseline data obtained across the three weeks prior to the beginning of releases. The corresponding 95% confidence intervals (CIs) were calculated by a 10,000-loop bootstrap for each period [25]. The CIs were calculated for the entire period of releases and for each period of seven weeks. The CIs were established in R version 3.5.2 (2018-12-20) (Copyright © 2018 The R foundation for Statistical Computing).

Analysis of Covariance (ANCOVA) using R version 3.5.2 was performed to verify the difference of viable progenies between control and treated areas over the study period. For this 29 weekly means of viable progeny by trap for both areas were calculated in the period when releases occurred.

Dengue incidence from March to May 2019 ((number of Dengue cases/exposed population of the area) X 100,000) were calculated for control (I_DC_) and treated (I_DT_) areas. The rate ratio (RR) is the ratio between the two incidences and was calculated by using the formula RR=I_DT_/I_DC_. Values of RR<1 indicate that the intervention (in this case, NVC treatment) is protective against Dengue and values of RR>1 indicate that intervention is a worsening factor for Dengue. Confidence intervals 95% were established in R software. ANCOVA was also used to analyze the differences between Dengue cases originated in control and treated areas.

To support the correlation between dengue cases, the mean of eggs and larvae progeny in each area, a linear regression was performed. Correlation was established based on the accumulated dengue cases that occurred in each area, compared to the sum of weekly mean of eggs collected and the weekly mean of viable larvae, to compare this variable, the number of dengue cases was paired with the data of eggs or viable larvae from 3 weeks before the dengue case was registered. This analysis was performed from the first week after the NVC releases started in the treated area and proceed until the end of field surveillance.

In addition, to better evaluate the influence of treatment and the number of viable larvae progenies on the total number of cases during the NVC releases, a Multivariate Anova was performed in R software. Both study areas were split to the three main neighborhoods in each region. Based on the 29 weeks of NVC releases, a mean of viable progeny per trap for each neighborhood was established and the total number of dengue cases for each neighborhood was calculated. Results were considered significant when p values were smaller than 0.05.

## Results

### Sterility of NVC male mosquitoes does not affect their competitiveness capacity

One of the problems of SIT is related to the method of sterilization of male insects, since several of the available techniques potentially affects the fitness of mosquitos and compromises their competitiveness capacity. To evaluate the impact of sterilization in NVC males, laboratory and semi-field tests were conducted to demonstrate the sterility and competitiveness of NVC mosquitoes. The laboratory-scale fertility bioassay using NVC male mosquitoes showed that they are unable to generate viable offspring after copulating with virgin females reared under similar conditions (Figure 2A and 2B). Although these results suggested that NVC are sterile, it is possible that this outcome is due to an inability of male mosquitoes to mate with females. Therefore, additional tests were performed to evaluate the ability of NVC to copulate and compete with normal fertile mosquitoes. As shown in Figure 2C and 2D, NVC male mosquitoes are able to compete with wild males when subjected to competitive assays using different ratios of sterile males to fertile males. To demonstrate that NVC mosquitoes were also competitive when exposed to field environmental conditions, a semi-field trial was conducted in the city of Jacarezinho. The results show that NVC mosquitoes were equally competitive with non-sterile mosquitoes under semi-field environmental conditions and proportionally suppressed the viable progeny of the next generation (Figure 2E and 2F).

**Figure 2.**
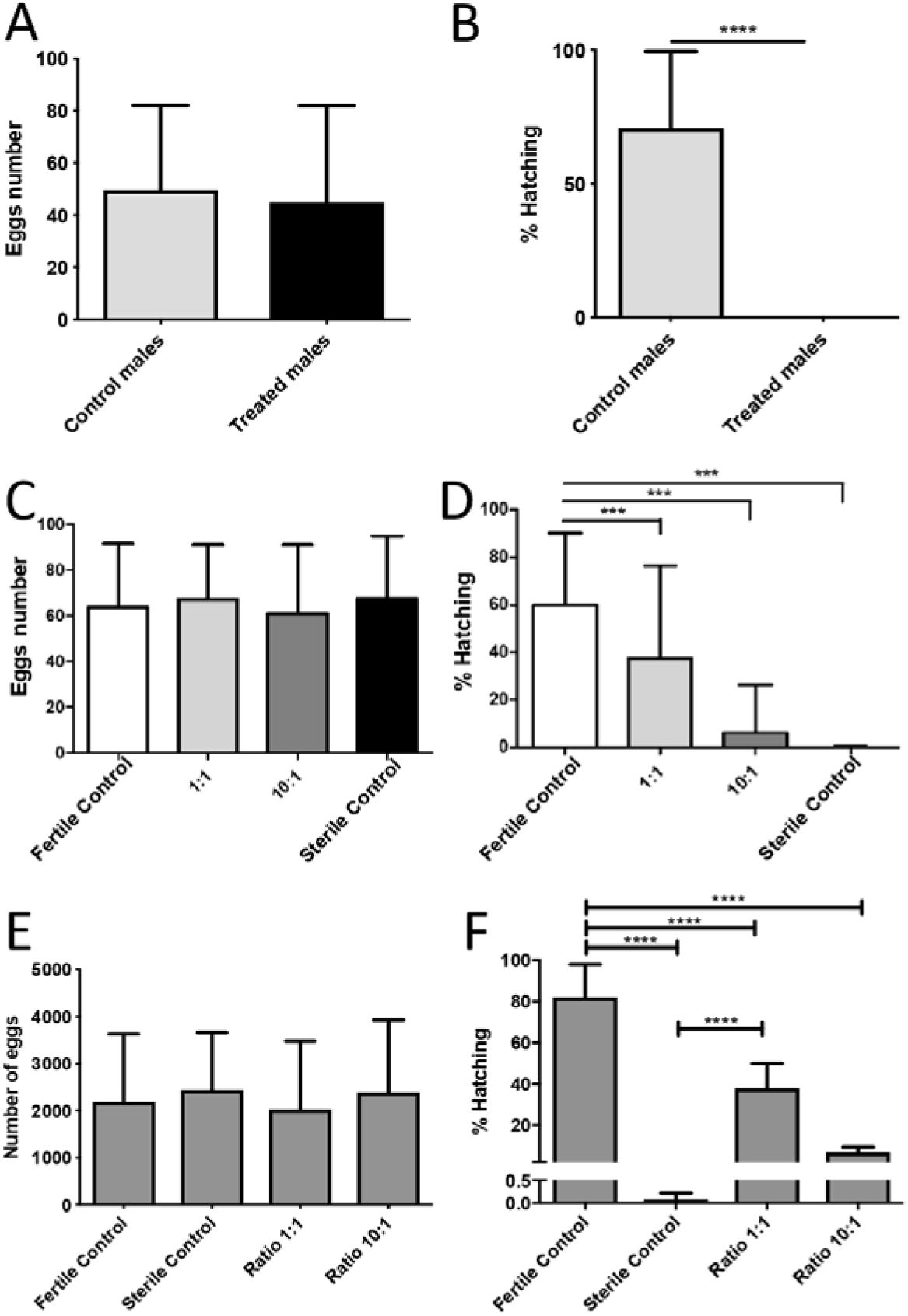
Effect of sterility treatment on viable *Ae. aegypti* progeny and competitive capacity of NVC mosquitoes. **A and B**. Seven-days-old NVC (Treated males) or normal fertile males (Control males) were allowed to mate with regular virgin females reared under similar laboratory conditions, for three days. The females were blood fed and allowed to oviposit their eggs in individual oviposition cages. Subsequently, the eggs were counted and hatched, in order to determine the percentage of viable larvae. In Panel A the average number of eggs per female is shown, and Panel B represents the percentage mean of viable larvae that hatched from the eggs. Data representative of 2 independent experiments. Statistical analysis: Unpaired t test, **** p <0.0001. **C and D**. Competitive capacity of NVC: Seven-days-old NVC were mixed with different ratios of fertile males (only untreated males – fertile control; 1 NVC: 1 untreated male; 10 NVC: 1 untreated male and only NVC – sterile control) and allowed to mate with regular virgin females reared under similar laboratory conditions. Blood feeding, oviposition and hatching was performed as described in A. Panel C: the average number of eggs per female, and Panel D represents the percentage average of viable larvae that hatched from the eggs. Data on 2 independent experiments. Statistical analysis: One-way Anova, Tukey test, *** p <0.005. **E and F**. Competitive competence of NVC in the semi-field trial. Different proportions of NVC and fertile male mosquitoes were placed in cages installed in a safe area in the city of Jacarezinho and allowed to mate with normal (fertile) virgin females, according to the protocol described in the Methods section. After the mate period, females from all groups were fed with blood and allowed to oviposit in appropriate containers placed inside the semi-field cages. The eggs were then removed from the cage, counted and hatched. E. Eggs average obtained in each experimental group from three independent experiments. F. Percentage of hatching (viable larvae derived from eggs) for each group (three experiments). Statistical analysis: One-way Anova (Tukey multiple comparison test, **** p <0.0001.

### Large-scale releases of NVC male mosquitoes successfully suppressed a field *Ae. aegypti* mosquito population

The field study was carried out to demonstrate the efficacy of the method in suppressing the wild mosquito population.

Community outreach was started in Jacarezinho already back in 2017 (appendix S5), and NVC mosquitoes were released starting from September 2018 to mid-April 2019. As already mentioned, this city was chosen, among other reasons, for presenting a history of dengue epidemics. The abundance of *Ae. aegypti* in Jacarezinho was monitored throughout 2017 and early 2018. As expected, peaks of mosquito infestation were during the hottest and wettest months of the year (from November to March) (Figure 3A).

**Figure 3.**
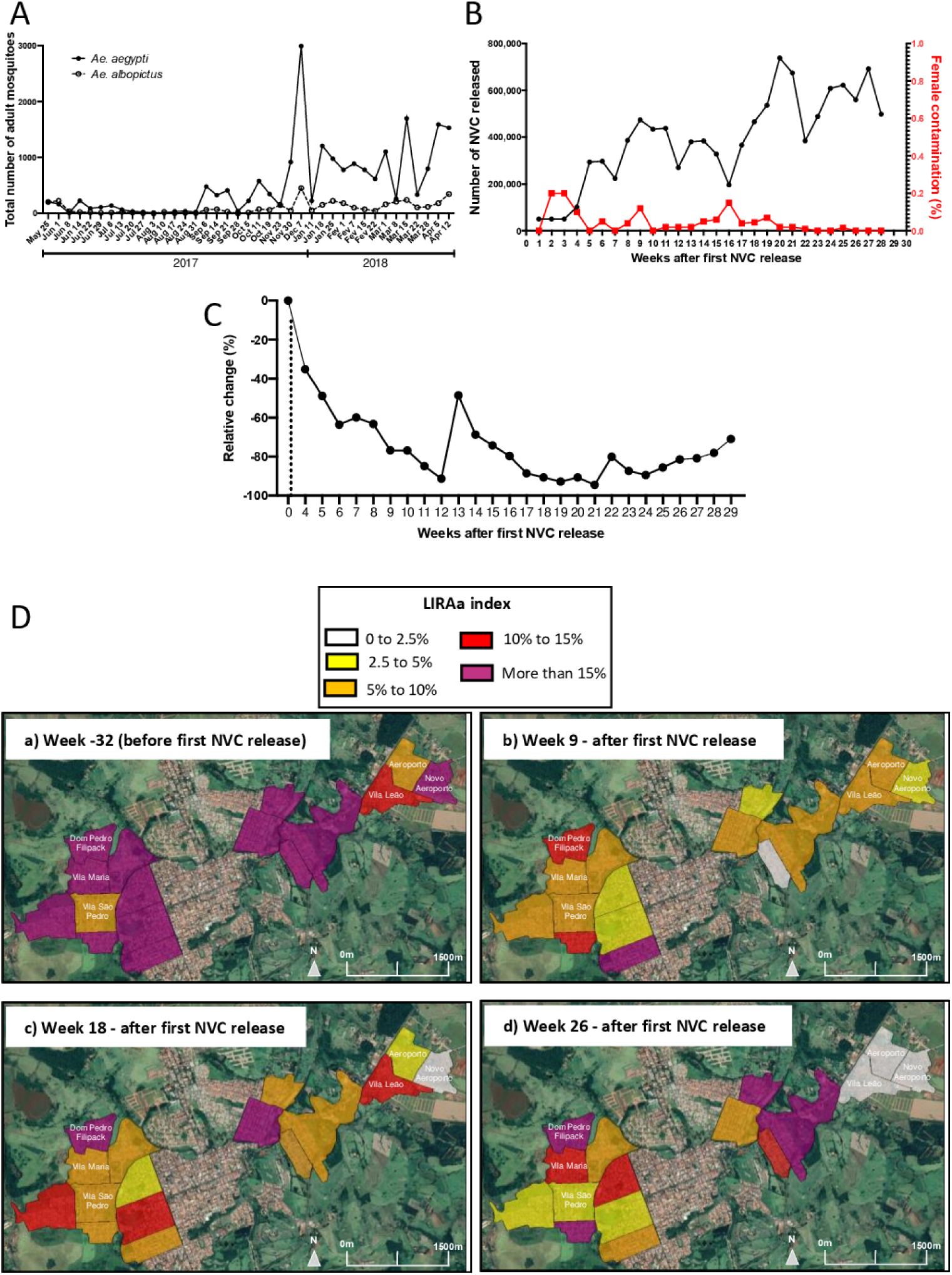
Massive releases of NVC male mosquitoes suppressed the local *Ae. Aegypti* population. **A**. Abundance of mosquito population *Ae. aegypti* and *Ae. albopictus* in the city of Jacarezinho from May 2017 until April 2018. Ovitraps were installed in 52 points distributed throughout Jacarezinho town and replaced weekly. The eggs from each ovitrap were hatched together and larvae were reared to adulthood. Adult mosquitoes were then identified by species. The data show the total number of adult mosquitoes of each species per collection. **B**. Number of NVC mosquitoes released at the treated area of Jacarezinho per week. The left Y axis (black) shows the absolute number of NVC mosquitoes released each week over the study period, while the right Y axis (red) shows the percentage of females present in the equivalent NVC release batch. **C**. Suppression of *Ae. aegypti* wild population in Jacarezinho after treatment with NVC mosquitoes. Weekly moving averages showing percentage change in *Ae. aegypti* abundance at the treated area, measured by mean number of larvae per trap relative to control area. **D**. Map of *Ae. aegypti* building infestation indices based on results of *Ae. aegypti* Infestation Index Rapid Survey (LIR*Aa*) by neighborhood. *Ae. agypt*i infestation before the implementation of NVC program is shown in upper left panel **(a)**, and infestation 9 **(b)**, 18 **(c)** and 26 **(d)** weeks after the beginning of NVC releases. Data were provided by Epidemiological Surveillance of the Health Department of Paraná. Municipality of Jacarezinho, Paraná State, Brazil.

The total number of NVC male mosquitoes released during the intervention period was calculated to be 12,335,200 and the number of NVC male mosquitoes used in each release is shown in Figure 3B. It is important to emphasize that the intention is to release only males, and not female mosquitoes, since it is the females that bite and potentially potentiate the spread of dengue and/or other arboviruses. Even though female mosquitoes reared in Forrest Innovations’ facility are pathogen-free, if released in enough numbers in an area where there is an ongoing epidemic, they could potentially contribute to local disease transmission. Therefore, each batch of sterile males underwent a quality control to detect the potential presence of contaminating females (Figure 3B).

Furthermore, to verify that NVC male mosquitoes remain viable and competitive after being released in the field, BG traps were installed in control and treated areas to recapture adult mosquitoes, including the NVC male mosquitoes. Although it is not possible to visually differentiate NVC mosquitoes from wild males, the fertility status showed that a high percentage of recaptured males was sterile, indicating that NVC mosquitoes remain viable, yet sterile, in the field at least one week after release (appendix S6).

To quantify the suppression effect of NVC male mosquito releases on the *Ae. aegypti* field population we used the method of weekly moving averages [25]. According to the established confidence intervals, weekly moving averages showed that treatment with NVC male mosquitoes in the treated area reduced the field population up to 91.4% (week 21 after beginning of NVC releases) compared to the mosquito population of the control area (Figure 3C). Our results on suppression of the local mosquito population are in accordance with the official *Ae. aegypti* infestation data provided by sanitary surveillance authorities. This survey is carried out continuously by the Ministry of Health of all Brazilian cities and is based on LIRAa index [24]. LIRAa index expresses the percentage of buildings that were positive for the presence *Ae. aegypti* larvae. As can be seen in Figure 3D, which indicates the LIRAa index in some neighborhoods of Jacarezinho, high *Ae. aegypti* infestation rates were found in the both control and treated areas before implementation of the NVC male mosquito release program. Most of these neighborhoods presented LIRAa index above 5%, and in some of them LIRAa was above 15%. As defined by the Brazilian Ministry of Health, an index above 4% classifies the monitored area as being at imminent risk of a dengue outbreak. Remarkably, 6 months after the beginning of the NVC releases, the treated area presented a drastic decrease in the rates of infestation of *Ae. aegypti* (from more than 15% to 0 – 2·5%), which placed these neighborhoods in a classification of a “satisfactory” situation. On the other hand, in neighborhoods of the control area, the LIRAa index continued to be extremely high and indicative of a risk of an imminent dengue outbreak, which indeed manifested in early March 2019. It is noteworthy that the neighborhoods immediately adjacent to the treated area also presented a high LIRAa index (more than 15%).

### Suppression of *Ae. aegypti* mosquito population by NVC males was associated with lower incidence of dengue in the treated area

Both treated and control areas presented a high incidence of dengue cases in 2010, 2011 and 2015 (Figure 4 A-C). In early March 2019 an outbreak of dengue began in Jacarezinho. During the period of NVC releases (from week 1 to 29, equivalent to October 3^rd^, 2018 and April 17, 2019, respectively), 293 confirmed dengue cases were reported in the entire city, and from this total, 109 cases (37.2%) originated in neighborhoods of the control area. In contrast, only 8 cases of dengue (2.7%) were reported in the neighborhoods treated with NVC male mosquitoes (Figure 4, panels D and D’). The clustering of dengue cases in several locations in the control area, as well as the rapid weekly increase is indicative of a high level of local transmission by mosquitoes. In contrast, the 8 cases reported in the treated area were sporadic and static during the period of NVC releases. On week 35, six weeks after the end of NVC male mosquitoes’ releases (Figure 4, panel E and E’) the accumulated number of dengue cases originated in the treated area remained much lower (16 cases) when compared to the control area (198 cases). As described in the Methods section, a blood sample for all the patients was collected to confirm the infection with dengue virus, both through serological and molecular analysis (RT-PCR Multiplex). The list of dengue cases based on their registered place of residence until the end of May, 2019 (week 35) can be found in appendix S7. In fact, the incidence of dengue in the control area during the entire period analyzed, from week 1 to 35 was 4,360 cases per 100,000 inhabitants, while the incidence of dengue in the treated area, in the same period, was only 264 cases per 100,000 inhabitants, which is almost 16 times lower than the dengue incidence in the control area. The Rate Ratio, which is the ratio between dengue incidence of the treated area and the control area, was 0.0606 (95% CI 0.0364-0.1006), indicating that the treatment with NVC had a protective effect for residents of the treated area in terms of dengue. Corroborating this, ANCOVA analysis showed that the difference between dengue cases in treated and control areas was statistically significant (appendix S8).

**Figure 4.**
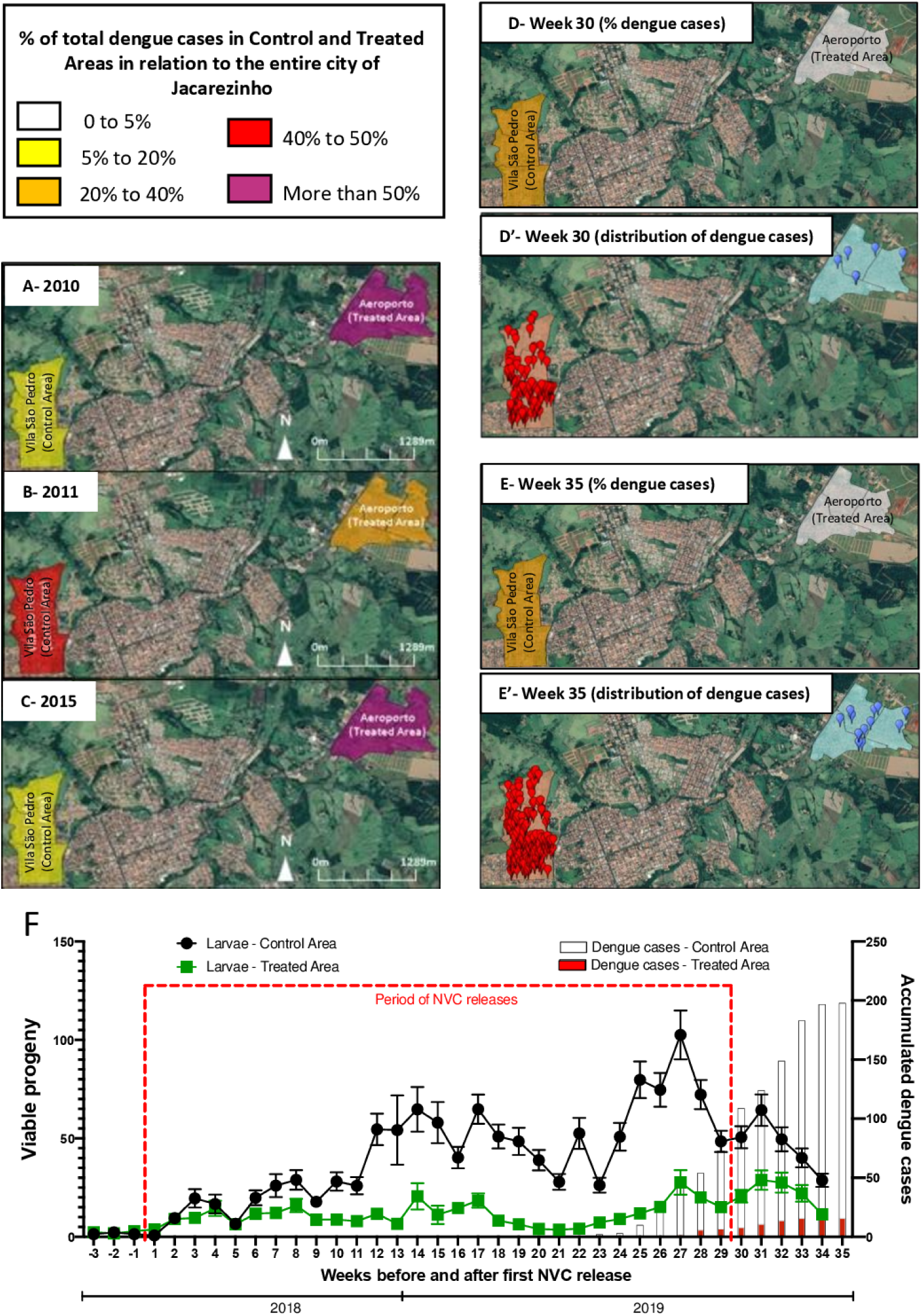
Distribution of dengue cases in the treated area and control of the city of Jacarezinho. Maps **A** (2010), **B** (2011) and **C** (2015) refer to the period prior to the start of treatment with NVC. Maps **D** and **D’** shows, respectively, the percentage of dengue cases and their distribution in control and treated area on week 30 (April 24, 2019), approximately 7 months after the start of NVC releases. Maps **E** and **E’** show, respectively, the percentage of dengue cases and their distribution in control and treated areas in Jacarezinho on week 35(May 28, 2019), six weeks after the end of the NVC release period. On maps **D’** and **E’**, blue marks indicate the points where dengue cases where reported in the treated area and red marks the dengue cases reported in the control area. Data were provided by Epidemiological Surveillance of the Health Department of Paraná. Municipality of Jacarezinho, Paraná state, Brazil. **F**. Mean viable progeny per ovitrap and dengue cases in control and treated areas overtime. Control and treated areas were monitored through egg collection (ovitraps) over 37 weeks. Releases of NVC in the treated area occurred between September 28, 2018 and April 22, 2019 (weeks 1 to 29), on a weekly basis. Eggs collected from the control and treated areas were transferred to the laboratory, where they were hatched. The mean number of larvae derived from eggs of each ovitrap (100 for each area) over the 37 weeks is represented in left-Y axis and was defined as viable progeny. Statistical analysis: Analysis of Covariance provided a p-value <0·0001 for the difference between slopes for mean larvae per trap from control and treated areas. The cumulative number of confirmed cases of dengue in the control area (gray bars) and treated (red bars) were provided by the Parana Health Department and are shown on the Y axis on the right. The incidence of dengue in control area (I_DC_) was 4·36% (95% CI 3·80%-4·99%) and in treated area (I_DT_) was 0·26% (95% CI 0·16%-0·43%). The Rate Ratio (I_DT_/ I_DC_ is 0·0606 (95% CI 0·0364-0·1006). Both Linear Regression and Multivariate Anova (correlation between viable progeny in treated and control areas and dengue cases) provided p-values < 0·05.

Finally, two different statistical analysis were performed to demonstrate the direct correlation between the low incidence of dengue and the significant reduction in viable progeny of *Ae. aegypti* mosquitoes overtime, which resulted from the release of NVC male mosquitoes (Figure 4F). First, a linear regression analysis showed that the increase in the number of dengue cases is related both to the mean of eggs collected and viable larvae per week. For both study areas significant p values were obtained, in the control area p=4.551e-06 and p=2.496e-06 for eggs collected and viable larvae, respectively. In the treated area p=1.824e-05 and p=9.058e-07 for eggs collected and viable larvae mean, respectively, evidencing the dependence between these variables (appendix S8). Then, a Multivariate Analysis (MANOVA) was performed to confirm this correlation (appendix S8). This analysis was based on mean number of viable larvae from eggs collected in the control and the treated areas during NVC releases and the number of dengue cases that originated in both areas in this period (Figure 4F). The analysis showed a strong influence of NVC treatment in reducing dengue cases (p value = 0.0025), which corroborates that the NVC releases has the potential to prevent dengue outbreaks (appendix S8).

## Discussion

Jacarezinho is a town with about 40,000 inhabitants. It experienced several dengue outbreaks in the last decade, most notably in 2010, 2011 and 2015. The geographic distribution of dengue cases in those past epidemics shows that both the control and treated areas presented the greatest number of cases in the town, and that overall, the proportion of cases in the treated area was slightly higher (Figure 4A-C). We set out to perform our study by targeting these neighborhoods for the release of the NVC sterile mosquitoes starting in September 2018 (week 1). As is common in such real-world circumstances [13], it was not logistically possible to use statistical methods to predetermine sample sizes, to randomize the experiments or to blind the investigators.

The first case of dengue in Jacarezinho was confirmed in the beginning of March 2019 (week 23) and the total number of cases increased until April 24^th^ 2019 (week 29, end of the NVC male mosquito release period) with 293 confirmed dengue cases. Out of these, 109 cases were reported in the control region, whereas only 8 cases were reported in the treated area. This difference of more than 90% between the number of dengue cases in the treated and control areas continued even 6 weeks after the total cessation of NVC male mosquito releases and demonstrates the success of our intervention program. During the last weeks of NVC male mosquito’ releases (weeks 28 and 29), the formal risk of dengue epidemic in Jacarezinho was declared by the local authorities, and by the end of May 2019 (week 35), a formal epidemic was confirmed, with 586 cases reported in total, 198 of which were in the control region (similar proportion of cases to past epidemics), whereas the treated region had only 16 cases (dramatically lower incidence compared with past epidemics). The details of the cases distribution are provided in appendix S7.

The data we present herein shows that an effective integrated SIT intervention plan can dramatically reduce the mosquito infestation *of Ae. aegypti*. Strikingly, the mosquito infestation index of the treated neighborhoods remained very low even though the directly adjacent neighborhoods were found to have very high mosquito indices in the LIRA survey. This underscores the robust nature of our NVC male mosquito intervention, since past studies have found significantly diminished effects of SIT in the “fringe areas” [13,15]. As such, allowing for such potential migration our suppression results become even more remarkable.

Critically, we have provided the first evidence that this new environmentally benign vector control intervention can potentially thwart the spread of a mosquito-borne disease epidemic. Our approach is based on the use sterile male mosquitoes (NVC) that are produced from a local mosquito strain and close collaboration with the local community and authorities.

With a dengue outbreak in the making, the local authorities could not ethically leave the control region without acting. Towards the end of March 2019, they began intervention in a series of reactive actions to try to thwart the outbreak and prevent an epidemic, including massive spraying of the organophosphate Malathion and ‘blocking’ (an intervention of seeking out larvae in breeding sites in a 100-meter radius around the infected case) of every resident dengue-case reported. This may have impacted the results and led to a certain reduction of the adult and subsequent larvae population in the control region (Figure 4F, weeks 27-29). Indeed, the incidence of dengue in the control area may have been even graver had the authorities not intervened at all. Due to the near absence of dengue cases in the treated area, no such reactive intervention was required. Our study directly demonstrates that these highly common reactive measures that are utilized all over the world [26–29] were ineffective in preventing the continued spike in the number of dengue cases in the control region. Together, these data provide strong evidence that near prevention of dengue transmission in the treated area is exclusively due to NVC male mosquito releases.

Indeed, a recent review on management of mosquito resistance concluded that increases in insecticide resistance development in *Aedes* vectors as arbovirus epidemics proliferate underscore the urgency to create Insecticide Resistance Management (IRM) programs to maintain or recover vector control efficacy, and that when control strategies using insecticides are implemented, they should be systematically associated with noninsecticidal tools and, when possible, replaced by alternative tools to reduce the selection pressure on *Aedes* populations and limit the evolution of resistance [30]. Herein we provide a highly effective alternative tool to thwart the spread of Aedes resistance to insecticides.

With the lack of specific antiviral drugs to treat dengue as well as uncertainties about the efficacy and safety of the dengue vaccine [8,31], integrated vector control remains the main WHO recommendation for the near future [1]. It is thus striking that there is paucity of reliable evidence for the effectiveness of any alternative dengue vector control method. Specifically, of the plethora of SIT studies conducted to date, none was able to directly show a reduction in dengue incidence, and entomological indices alone were used as end points [11].

One of the key factors for successful SIT implementation for mosquito control is identifying a technology that induces sterility while at the same time retaining the sterilized treated males vigorous and capable of successfully competing with the endemic male population in the release area. Furthermore, in order to be implemented widely, such a method needs to be robust, cost-effective, easily scalable, and easy to implement in remote areas where it is most needed [32]. Unfortunately, all the existing biological control alternatives have multiple limitations that make them unavailable for immediate globally impactful implementation (Table 2). For example, RIDL mosquitoes have recently been shown not be self-limited, and surviving males have integrated their gene legacy into the local population [16], thus raising additional concerns beyond those that have prevented this intervention from being implemented. Indeed, 10 years since it was first tested in the Cayman Islands [33], there is still great difficulty in bypassing negative public sentiment to test it in the Florida Keys, and its operation in Brazil has almost ground to a halt. Population replacement strategies, such as the “Eliminate Dengue” program, based on *Wolbachia* inhibiting replication of viruses, showed some success in Australia [34], but scaling up this strategy seems daunting since there are multiple challenges to address such as the potential of the viruses to overcome *Wolbachia*-mediated “blocking” [17,35], reduced fitness of multiple strain *Wolbachia* carrying mosquitoes [17,36–38], and potential of high temperatures in some locales to constrain the ability of *Wolbachia* to invade natural mosquito populations and block disease transmission [17,38]. Recently, a combined IIT/SIT approach was hailed to almost eliminate mosquitoes using a *Wolbachia*-male + irradiated female contamination combination to facilitate population suppression. Notwithstanding the small size of the intervention areas and the large number of mosquitoes per area due to their reduced competitiveness, this method is very difficult to implement on a large scale, requiring backcrossing to local populations everywhere that it is implemented. Moreover, even under the very stringent conditions of the trial, several WPi bearing females were recovered, underscoring the potential of limited population suppression inadvertently becoming population replacement once the scope of this kind of intervention is enlarged [13]. This is so because quality control for successful irradiation to the females is inevitably completed only after the batch was released.

**Table 2.** Comparative analysis of the different methods for Vector control based on SIT, IIT or “blocking”.

| Method | *SIT* (irradiation) | *Wolbachia* (IIT) | *Wolbachia* + irradiation (IIT/SIT) | *Wolbachia* (Transinfection - TI) | Release of Insects carrying a Dominant Lethal (RIDL) | Natural Vector Control |
| --- | --- | --- | --- | --- | --- | --- |
| **Details** | Release of irradiated-male mosquitoes | Release of *Wolbachia*-infected males (Incompatible Insect Technique - IIT) | Combined IIT/SIT (irradiated mosquitoes) | Release of males and females infected with *Wolbachia* (Blocking of virus transmission) | Release of transgenic mosquitoes (OX513A) | Release of NVC sterile male mosquitoes |
| **Field trial** | Bellini et al., 2013[39] | Mains et al., 2019[15] | Zheng et al., 2019[13] | Hoffmann et al., 2011[34] | Carvalho et al. 2015[14] | Present Study |
| **City** | Emilia-Romagna region (five small towns), northern Italy | South Miami, FL, USA | Guanzhou, China | Cairns, Australia | Juazeiro, Bahia, Brazil | Jacarezinho, Parana, Brazil |
| **Treated Area** | 112 ha | 170 ha | 32.5 ha | Not Informed | 11 ha | 77 ha |
| **Period of releases** | 3 years | 6 months | 2 years | 10 weeks |  | 7 months |
| **Mosquitoes released** | 2 M | 6.8 M | 197.1M | 300,000 adults | 185,000 males | 12.3M |
| **Field monitoring** | 85 ovitraps | 70 ovitraps and 35 BGs | 150 ovitraps and 60 BGs | 320 ovitraps | 120 ovitraps and BGs (amount not informed) | 100 ovitraps and 20 BGs |
| ***Ae. aegypti* population reduction relative to control** | Induced up to 68% sterility in the native population (eggs' hatching %). No direct demonstration of suppression. | Up to 78% in the center<br>Max 45% in the "edge" areas | Up to 94% | Successful fixation of *Wolbachia* in local mosquito population occurred after 11 weeks of releases | Successful suppression of field population (81-95%) | Up to 91% throughout |
| **% female contamination** | Not informed | Not informed | 0.6% | Not applicable | 0.02% | 0.003% |
| **Reduction in the number of dengue cases** | Not evaluated | Not evaluated | Not evaluated | Not evaluated | Not evaluated | 93% less (220 cases in the control area and only 16 cases in treated area – until June 2019) |
| **Risk of introduction of new organisms/ genes/ residues into the nature** | Low efficiency of sterilization by irradiation results in the release of large numbers of fertile males | High (undesired *Wolbachia* introduction into natural mosquito populations [17,40]) | Medium (sterilization by irradiation did not prevented completely undesired introduction of *Wolbachia* into natural population) [13] | High (introduction of *Wolbachia* into natural mosquito population is the basis of this method) | High (not self-limited: spread of transgene to natural population [16]) | No risk |
| **Impact on mosquito fitness or competitive capacity** | High | High, depending on the *Wolbachia* strain [17,41,42] | High [43] | High, depending on the *Wolbachia* strain [17] | High [44] | No impact, sterile males are produced from local mosquito population |
| **Long-term efficacy** | Need to perform additional releases in the subsequent mosquito seasons | Need to perform additional releases in the subsequent mosquito seasons | Need to perform additional releases in the subsequent mosquito seasons | Need to periodically check for the presence of *Wolbachia* in field-recaptured mosquitoes from field [17] | Need to perform additional releases in the subsequent mosquito seasons | Need to perform additional releases in the subsequent mosquito seasons |
| **Additional comments** | Because irradiated males presented lower competitiveness capacity they are not the first choice for SIT programs. | Poor competitive capacity implies the need to release greater amounts of males<br>High cost program (reviewed in [17]) | Poor competitive capacity implies the need to release greater amounts of males<br>High cost program: difficult to implement in large-scale due the irradiation process [17,43] | Potential loss of infected mosquito population decreases the effectiveness of virus blocking (reviewed in [17])<br>Risk of selecting resistant and more virulent strain of virus [17,45]<br>Evolutionary changes in host genome resulting in reduced virus blocking even in the presence of *Wolbachia* [17] | Historic public rejection, lack of support from authorities, long and difficult regulatory process associated to use of GM mosquitoes and the recently shown gene flow from GM mosquitoes into the local gene pool [16] | Only one field trial performed so far |
| **Conclusions** | Medium efficacy. Recently this technique has been used in combination with other techniques, which involves additional costs for the program.[13,46] | Medium efficacy demonstrated, however the risk of undesired introduction of *Wolbachia* is a strong limitation of the method. | High efficacy when males are released in great amounts | Effective at least in a short-term, but effects of introduction of *Wolbachia* at long-term is not known so far. | Although mosquito suppression is effective, the potential environmental impact, the history of public rejection and lack of support from the authorities difficult implementation of this method. | High effective reduction in the natural mosquito population; Dramatic reduction in number of dengue cases. No environmental impact: self-limited mosquitoes; no risk of introduction of new mosquito strains, genes, residues into environment. |

The process described herein to induce male sterility in *Ae. aegypti* overcomes all the technological, logistical and ecological hurdles described above, as well as successfully addressing regulatory compliance and the public concerns. We developed a treatment that is transient in nature and the active ingredients do not persist in the released adult mosquito [47] (appendix S9). By collecting thousands of local mosquito eggs before the start of the project, we were able to use the ambient mosquito genepool to guarantee that the local climate adaptation and local female seeking capabilities of the released sterile males will be optimal. By creating a colony with hundreds of “founders” and thus large genetic variability, we were able to ensure that the males were highly competitive and survived at least one week after their release.

In addition, as has been determined in previous studies, the education and involvement of the community in decision making provides a crucial component in successful implementation of SIT programs. A bottom-up approach that targets school workshops, community programs and total transparency, was highly successful in exposing the people to the benefits of releasing sterile male mosquitoes (appendix S5).

Dengue outbreaks are highly unpredictable, but it has been empirically determined in several previous studies that a minimum mosquito-vector threshold is needed to facilitate the spread of the virus in the population. Gradual mosquito population suppression all the way to over 91·4% reduction from the control area that was demonstrated in this study shows the importance of sustaining a continuous release program that is dependent on real-time monitoring of the mosquito population. Concurrent programs run over large regions may well bring total relief from *Aedes*-related diseases by the second year of implementation.

The dramatic influence on the incidence of malaria by the deployment of Pyrethroid-treated bed nets underscores the ability of the international health community and Non-Governmental Organizations to mobilize in order to take advantage of an effective system to reduce disease. However, these efforts require the massive integrated coordinated action and funding of nations and public organizations such as the Bill and Melinda Gates Foundation in order to be effectively disseminated. In order to create real impact on a global scale, local communities need to be empowered to set up the infrastructure for them to be able to endorse the transformative paradigm that is described herein. It is our humble hope that this communication will be the trigger for such global mobilization to further test and subsequently employ NVC male mosquitoes wherever the threat of dengue and other arboviral diseases is significant.

## Data Availability

All relevant data are within the paper and its supporting information files.

## Supporting information captions

Appendix S1– Fertility bioassay

Appendix S2– Semi-field trial design

Appendix S3– Local Ethic Approval

Appendix S4– Agreement form for installation of ovitraps

Appendix S5– Community outreach and educational program

Appendix S6– Evaluation of NVC viability after release

Appendix S7– Dengue cases distribution

Appendix S8– Statistical analysis

Appendix S9– Residues analysis

